# Effect of SARS-CoV-2 infection on outcome of cancer patients: A systematic review and meta-analysis of studies of unvaccinated patients

**DOI:** 10.1101/2021.10.20.21265284

**Authors:** Giulia Di Felice, Giovanni Visci, Federica Teglia, Marco Angelini, Paolo Boffetta

**Affiliations:** Department of Medical and Surgical Sciences, University of Bologna, Bologna, Italy; Stony Brook Cancer Center, Stony Brook University, Stony Brook, NY, USA

**Keywords:** SARS-CoV-2, COVID-19, Cancer, Mortality, ICU, Disease severity, Meta-analysis

## Abstract

**Background:** Since the beginning of the SARS-Cov-2 pandemic, cancer patients affected by COVID-19 have been reported to experience poor prognosis; however, a detailed quantification of the effect of SARS-CoV-2 infection on outcome of unvaccinated cancer patients has not been performed.

**Methods:** To carry out a systematic review of the studies on outcome of unvaccinated cancer patients infected by Sars-Cov-2, a search string was devised which was used to identify relevant publications in PubMed up to December 31, 2020. We selected three outcomes: mortality, access to ICU, and COVID-19 severity or hospitalization. We considered results for all cancers combined as well as for specific cancers. We conducted random-effects meta-analyses of the results, overall and after stratification by region. We also performed sensitivity analyses according to quality score and assessed publication bias.

**Results:** For all cancer combined, the pooled odds ratio (OR) for mortality was 2.32 (95% confidence interval [CI] 1.82-2.94, I^2^ for heterogeneity 90.1%, 24 studies), that for ICU admission was 2.39 (95% CI 1.90-3.02, I^2^ 0.0%, 5 studies), that for disease severity or hospitalization was 2.08 (95% CI 1.60-2.72, I^2^ 92.1%, 15 studies). The pooled mortality OR for hematologic neoplasms was 2.14 (95% CI 1.87-2.44, I^2^ 20.8%, 8 studies). Data were insufficient to perform a meta-analysis for other cancers. In the mortality meta-analysis for all cancers, the pooled OR was higher for studies conducted in Asia than studies conducted in Europe or North America. There was no evidence of publication bias.

**Conclusions:** Our meta-analysis indicate a two-fold increased risk of adverse outcomes (mortality, ICU admission and severity of COVID-19) in unvaccinated cancer patients infected with SARS-CoV-2 compared to uninfected patients. These results should be compared with studies conducted in vaccinated patients; nonetheless, they argue for special effort to prevent SARS-CoV-2 infection in patients with cancer.

**Funding:** No external funding was obtained.

## Introduction

Since the emergence of SARS-CoV-2, many studies have been conducted on the outcomes of COVID-19, in order to identify factors associated with a higher death rate and a more severe infection course. Some groups of patients at increased risk of severe COVID-19, morbidity and mortality have been identified, including elderly patients, and those with comorbidities, as hypertension, diabetes, chronic kidney disease or COPD [1]. Cancer patients are also a high-risk group due to their compromised immune systems and vulnerability to infection resulting from their disease and treatments [2].

It is generally assumed that cancer patients are at higher risk for severe COVID-19 and death attributed to COVID-19 [3]. However, cancer encompasses a very heterogeneous group of diseases with a diverse range of subtypes and stages. In addition, not all cancers are equal in terms of incidence, prognosis, and treatment. This must be taken into account when the type of cancer is not specified [4]. For this reason, although descriptions and analyses of risk factors, clinical courses, and mortality in cancer patients infected with SARS-CoV-2 have been reported, a quantitative assessment of the effect of COVID-19 in patients with cancer would be important to guide clinical decision-making.

We aimed at conducting a systematic review of the epidemiological features of the studies of COVID-19 in cancer patients conducted before the implementation of vaccination campaigns, and to provide a quantitative estimate of the risk of cancer patients for severe infection course and COVID-19 mortality, compared to uninfected cancer patients. We decided to restrict our review to studies of unvaccinated patients because (i) they provide the clearest picture of the effect of SARS-CoV-2 infection on outcome of cancer patients, and (ii) the full effect of vaccination might not have been yet captured by available studies.

### Materials and methods

This systematic review was conducted according to the PRISMA statement [5]. We submitted the protocol (available as Supplementary File 1) to the PROSPERO Registry. To carry out the systematic review of the scientific literature, the following string was used for the PubMed database:

> *(neoplas*[TIAB] OR tumor*[TIAB] OR cancer*[TIAB] OR malignancy[TIAB]) AND (2019 novel coronavirus[TIAB] OR COVID-19[TIAB] OR COVID19[TIAB] OR SARS-CoV-2[TIAB] OR 2019-nCoV[TIAB])*.

In order restrict the review to studies populations on unvaccinated cancer patients, we included papers published in peer-reviewed journals up to December 31, 2020. We excluded abstracts and non-peer-reviewed reports, articles in languages other than English, and studies including children. We also excluded reviews, meta-analysis and case reports, and studies with less than 50 patients or less than 10 events. Finally, we excluded studies in which diagnosis of SARS-Cov-2 infection was not made by PCR testing.

The articles were independently reviewed and abstracted by 2 pairs of reviewers [GDF and MA; GV and FT], on the basis of title, abstract and full text; the disagreement between the authors of the reviews (6.1% of all studies) and was resolved through discussion with a fifth reviewer [PB].

We selected the following outcomes: mortality, ICU admission, severity of COVID-19 symptoms, and hospitalization: we combined these latter two outcomes because the definition of severity was heterogeneous across studies and the number of available studies was low. We excluded from the review studies addressing the impact of SARS-Cov-2 infection on prevention, diagnosis, and treatment of cancer patients, as well as studies on the oncogenic effect of the virus, e.g., analyses of cancer-related alterations. In addition, we carried out a back-search by inspecting the lists of references of articles selected for the review.

Figure 1 shows the flowchart for selection of the studies. Details on the studies retained in each step of the process are available from the authors.

**Fig 1.**
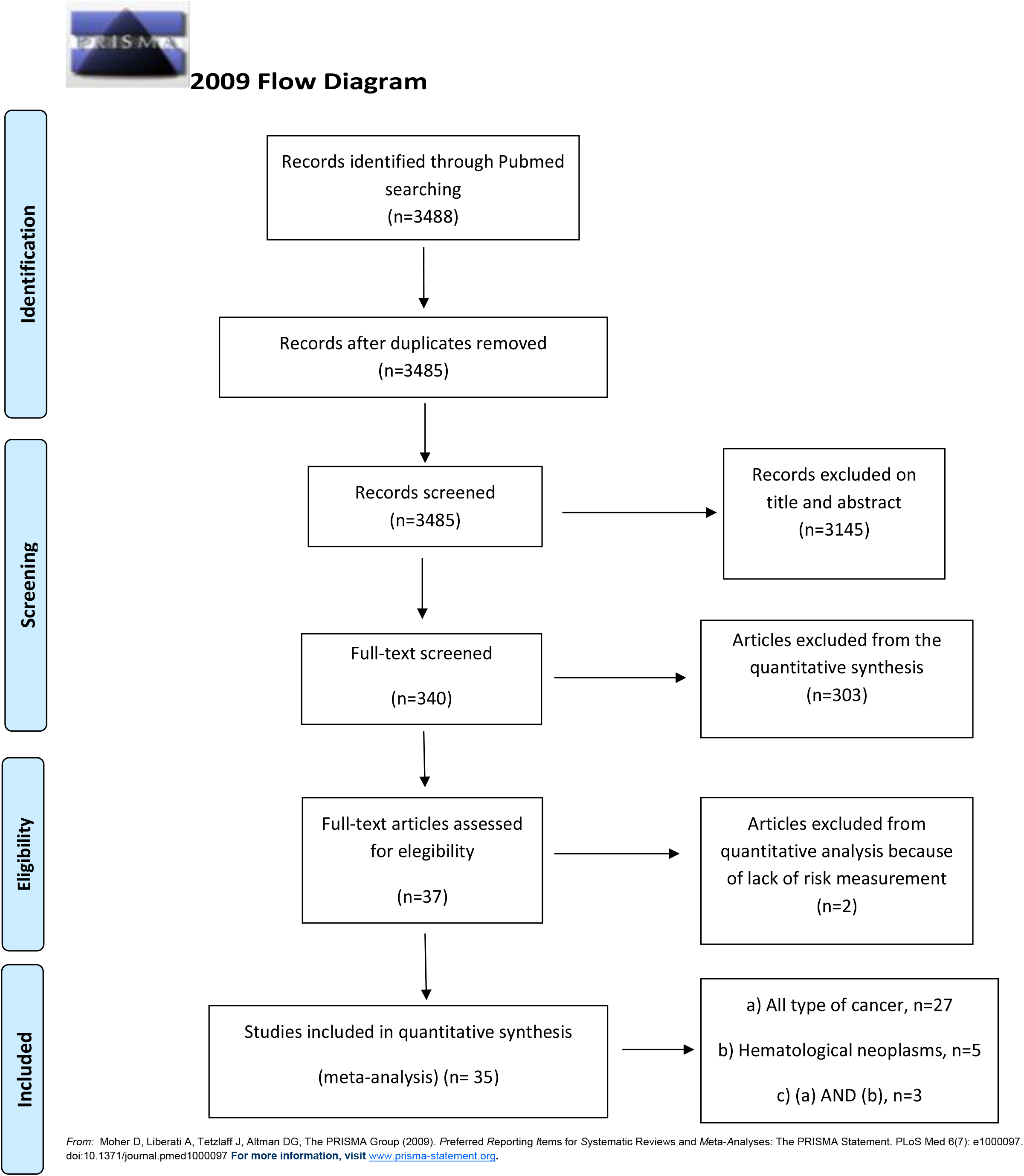
PRISMA

We abstracted the following parameters from the articles retained for the review: country, sample size, number of person affected by cancer and by SARS-Cov-2 infection, cancer type and comparison group (patients without cancer or patients with a different type of cancer), outcome, and risk estimate (relative risk or odds ratio [OR]) with 95% confidence interval (CI). If the risk estimate or the CI were not reported in the publication, we calculated them from the raw data, if possible. We also performed a quality assessment (QA) based on a modified version of CASP score [6], that included 10 criteria.

#### Statistical analysis

We conducted random-effects [7] meta-analyses of the risk estimates for the combinations of cancers and outcomes with more than five independent results. We also conducted stratified meta-analyses according to geographic region, to explore potential sources of heterogeneity, that we quantified using the I^2^ test [8].

To evaluated results stability, we performed sensitivity analyses by quality score and repeated the meta-analysis after excluding one study at a time. We also conducted secondary analyses excluding studies with results calculated on the basis of raw data. Furthermore, we considered the funnel plot and performed the Egger’s regression asymmetry test to assess publication bias [9].

Finally we conducted a cumulative meta-analysis, based on date of publication of subsequent studies.

Analyses were performed by STATA16 program [10], using specific commands metan, metabias, and metafunnel.

## Results

We identified a total of 3488 publications from the literature search, and excluded three because they were duplicates. We screened the titles and abstracts of 3485 articles: we excluded 3145 of them because not relevant (Figure 1), and retained 340 articles as potentially eligible.

After reviewing the full-texts, we excluded 303 articles because these did not meet the inclusion criteria, and included the remaining 37 studies in the review: we finally included 35 of them in the quantitative synthesis.

Among the 35 studies, 30 reported results for all cancers combined, and 8 for hematologic neoplasms (three of these reported both sets of results). Results for other specific cancers were sparse, and we could not conduct meta-analyses for them. Out of the 35 studies, 13 were from Europe, 11 from North America (all from USA), and 11 from Asia (9 from China and two from Iran). Fifteen studies were considered good quality (CASP score >9.5), nineteen studies were of moderate quality (9.5 ≥ CASP score > 6), whereas one was considered inadequate (CASP score ≤ 6).

Tables 1 and 2 show the details of the studies included in the analysis.

**Table 1.** Selected characteristics of studies included in the meta-analysis - All cancers

| Reference | Country | N patients | N outcomes | Quality score | Comparison | Outcome | RR/OR* | 95% CI |
| --- | --- | --- | --- | --- | --- | --- | --- | --- |
| Dai et al.-2020 [11] | China | 641 | 105 | 9.25 | Internal | Mortality | 2.34 | 1.15-4.77 |
|  |  |  |  |  |  | ICU | 2.84 | 1.59-5.08 |
|  |  |  |  |  |  | Severity | 2.79 | 1.75-4.44 |
| Haase et al.-2020 [12] | Denmark | 323 | 15 | 11.5 | Internal | Mortality | 3.18 | 1.66-6.09 |
| Meng et al.-2020 [13] | China | 436 | 109 | 9.5 | Internal | Mortality | 2.98° | 1.76-5.05 |
| Sun et al.-2020 [14] | USA | 323 | 67 | 10.5 | Internal | Mortality | 5.67° | 1.49-21.58 |
|  |  |  |  |  |  | ICU | 1.91° | 0.90-4.06 |
|  |  |  |  |  |  | Hospitalization | 2.16° | 1.12-4.17 |
| Nogueira et al.-2020 [15] | Portugal | 20293 | 611 | 9.25 | Internal | Mortality | 1.48 | 1.07-2.05 |
| Zandkarimi et al.-2020 [16] | Iran | 1831 | 32 | 6.5 | Internal | Mortality | 3.57 | 1.82-7.02 |
| Zhao et al.-2020 [17] | China | 539 | 23 | 6.5 | Internal | Mortality | 3.23 | 1.39-7.51 |
| Harrison et al.-2020 [18] | USA | 31461 | 1966 | 9.75 | Internal | Mortality | 0.87° | 0.72-1.09 |
| Gupta et al.-2020 [19] | USA | 2215 | 112 | 7.5 | Internal | Mortality | 2.20 | 1.50-3.22 |
| Ganatra et al.-<br>2020 [20] | USA | 2476 | 195 | 9.25 | Internal | Mortality | 3.53 | 2.95-4.23 |
|  |  |  |  |  |  | Severity | 3.75 | 3.17-4.44 |
|  |  |  |  |  |  | Hospitalization | 2.78 | 2.37-3.26 |
| Westblade et<br>al.-2020 [21] | USA | 2294 | 100 | 8.5 | Internal | Mortality | 1.29 | 1.04-1.61 |
|  |  |  |  |  |  | Severity | 0.76 | 0.57-1.01 |
| Wang et al.-<br>2020 [22] | USA | 16570 | 1200 | 11 | Internal | Mortality | 3.20 | 2.89-3.55 |
|  |  |  |  |  |  | Hospitalization | 2.85 | 2.63-3.09 |
| Cherri et al.-<br>2020 [23] | Italy | 2039 | 53 | 11 | Internal | Mortality | 2.22° | 1.25-3.94 |
| Gorgulu et al.-<br>2020 [24] | Turkey | 483 | 75 | 11 | Internal | Mortality | 1.81° | 0.88-3.72 |
|  |  |  |  |  |  | ICU | 2.14 | 1.26-3.63 |
| Jimenez et al.-<br>2020 [25] | Spain | 1549 | 103 | 9.75 | Internal | Mortality | 4.29° | 2.40-7.67 |
| Thompson et<br>al.-2020 [26] | UK | 470 | 87 | 10 | Internal | Mortality | 2.20° | 1.27-3.81 |
| Li et al.-2020<br>[27] | China | 1859 | 65 | 9 | External | Mortality | 1.59° | 0.94-2.68 |
| Shoumariyeh et<br>al.-2020 [28] | Germany | 78 | 39 | 8.5 | External | Mortality | 1.01 | 0.41-2.49 |
|  |  |  |  |  |  | Severity | 1.15 | 0.61-2.17 |
| Mehta et al.-<br>2020 [29] | USA | 1308 | 218 | 8.5 | External | Mortality | 2.38 | 1.69-3.35 |
| Rogado et al.-<br>2020 [30] | Spain | 42495 | 45 | 8 | External | Mortality | 4.82 | 2.67-8.71 |
| Brar et al.-2020 [31] | USA | 585 | 117 | 9.5 | External | Mortality | 0.98 | 0.58-1.66 |
|  |  |  |  |  |  | Severity | 0.80 | 0.57-1.13 |
| Zhang et al.-2020 [32] | China | 217 | 112 | 9.5 | External | Mortality | 4.83 | 2.87-8.12 |
|  |  |  |  |  |  | ICU | 2.60 | 1.87-3.62 |
|  |  |  |  |  |  | Severity | 1.43 | 1.09-1.87 |
| Sorouri et al.-2020 [33] | Iran | 159 | 53 | 10 | External | Mortality | 3.27° | 0.93-11.55 |
|  |  |  |  |  |  | ICU | 1.52° | 0.56-4.12 |
| Lunski et al.-2020 [34] | USA | 5145 | 312 | 9.5 | External | Mortality | 2.03° | 1.44-2.87 |
| Atalla et al.-2020 [35] | USA | 339 | 27 | 10.5 | Internal | Hospitalization | 3.34 | 1.81-6.16 |
| Cheng et al.-2020 [36] | China | 1476 | 29 | 6 | Internal | Severity | 2.14 | 0.97-4.73 |
| Song et al.-2020 [37] | China | 961 | 21 | 7 | Internal | Severity | 2.77 | 1.16-6.62 |
| Liang et al.-2020 [38] | China | 1590 | 18 | 8.75 | Internal | Severity | 4.07° | 1.23-13.45 |
| Bauer et al.-2020 [39] | USA | 1449 | 108 | 8.5 | Internal | Severity | 1.72° | 1.11-2.67 |
| Tian et al.-2020 [40] | China | 751 | 232 | 9 | External | Severity | 3.75 | 2.71-5.19 |

RR: Relative Risk OR: Odds Ratio CI: Confidence interval
ICU: Intensive Care Unit
\* Results derived from data reported in the publication are in italics
°Risk estimates derived from multivariate analysis

**Table 2.** Selected characteristics of studies included in the meta-analysis - Haematological tumors

°Risk estimates derived from multivariate analysis
| AUTHOR – YEAR | COUNTRY | SAMPLE SIZE (n) | CANCER (n)<br>O number of | QUALITY<br>ASSESSMENT | COMPARISON | OUTCOME | RR/OR* | IC |
| --- | --- | --- | --- | --- | --- | --- | --- | --- |
| Dai et al.-2020<br>[11] | China | 641 | 9 | 9.25 | Internal | Mortality | 9.07 | 2.16-38.13 |
| Haase et al.-2020<br>[12] | Denmark | 323 | 13 | 11.5 | Internal | Mortality | 1.83 | 0.85-3.93 |
| Meng et al.-2020<br>[13] | China | 327 | 16 | 9.5 | Internal | Mortality | 2.83° | 0.96-8.32 |
| Ygenoglou et al.-2020<br>[41] | Turkey | 1480 | 740 | 10.5 | External | Mortality | 2.20 | 1.93-2.50 |
| Shah et al.-2020<br>[42] | UK | 1183 | 68 | 9.75 | External | Mortality | 1.74° | 1.12-2.71 |
| Sanchez-pina et al.-<br>2020<br>[43] | Spain | 92 | 39 | 10.25 | External | Mortality | 6.65° | 1.87-23.67 |
| Passamonti et al.-<br>2020<br>[44] | Italy |  | 536 | 11 | External | Mortality | 2.04° | 1.77-2.35 |
| Cattaneo et al.-2020<br>[45] | Italy | 204 | 102 | 9 | External | Mortality | 2.10 | 1.14-3.85 |

CASP: Critical Appraisal Skills Programme
RR: Relative Risk
OR: Odds Ratio
IC: Interval Confidence
ICU: Intensive Care Unit
\*italics characters when calculated manually

Figures 2-3-4 reports the results of the meta-analyses of studies of patients with all cancers combined compared to patients without cancer, for mortality, admission to ICU and hospitalization or severity of COVID-19, respectively. The pooled OR for mortality was 2.32 (95% CI 1.82-2.94, I^2^ 90.1%), that for ICU admission was 2.39 (95% CI 1.90-3.02, I^2^ 0.0%), and that for hospitalization/severity of disease was 2.08 (95% CI 1.60-2.72, I^2^ 92.1%).

**Fig 2.**
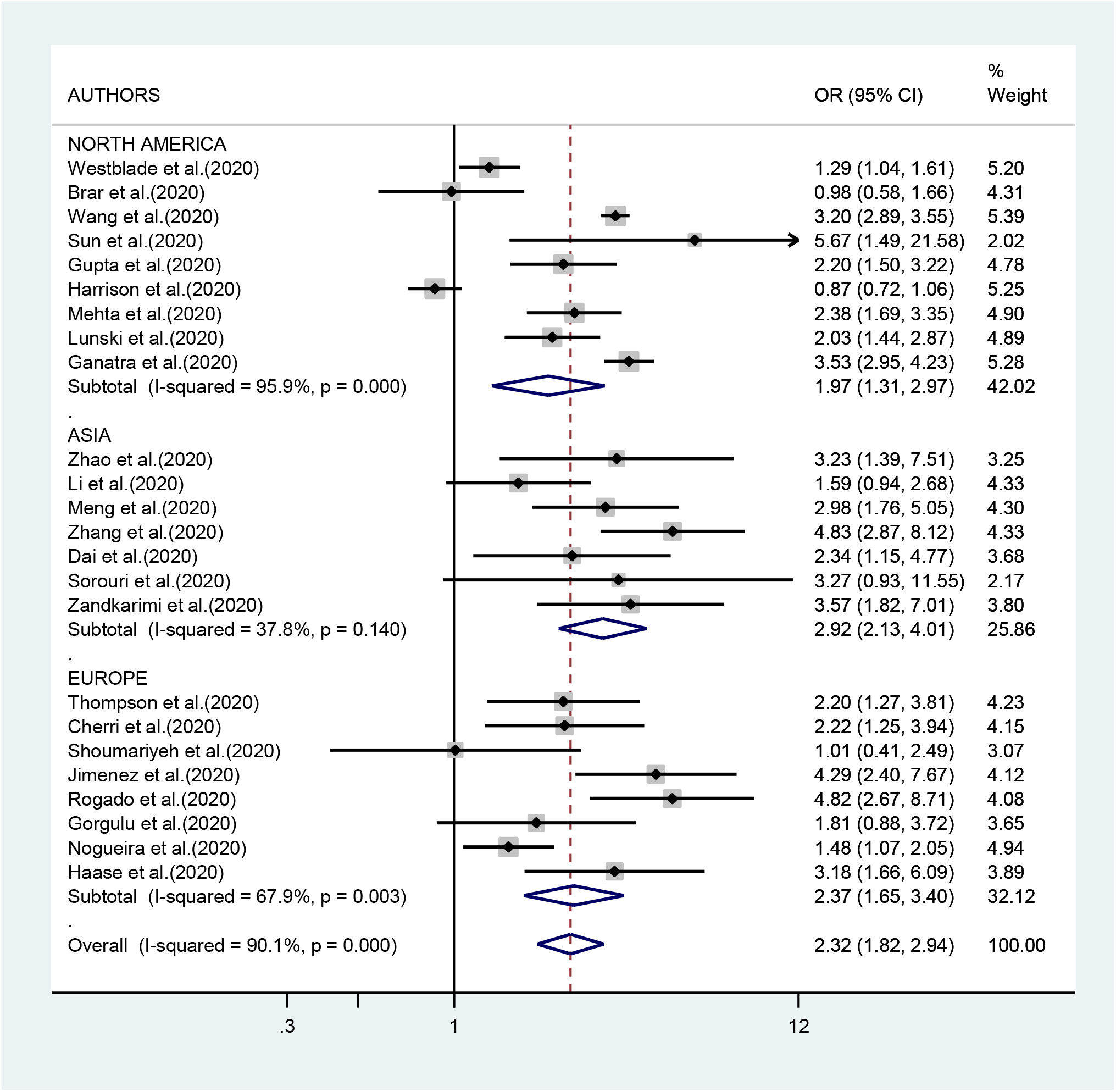
All type of cancer: Outcome 1 (Mortality) by Continental of publication

**Fig 3.**
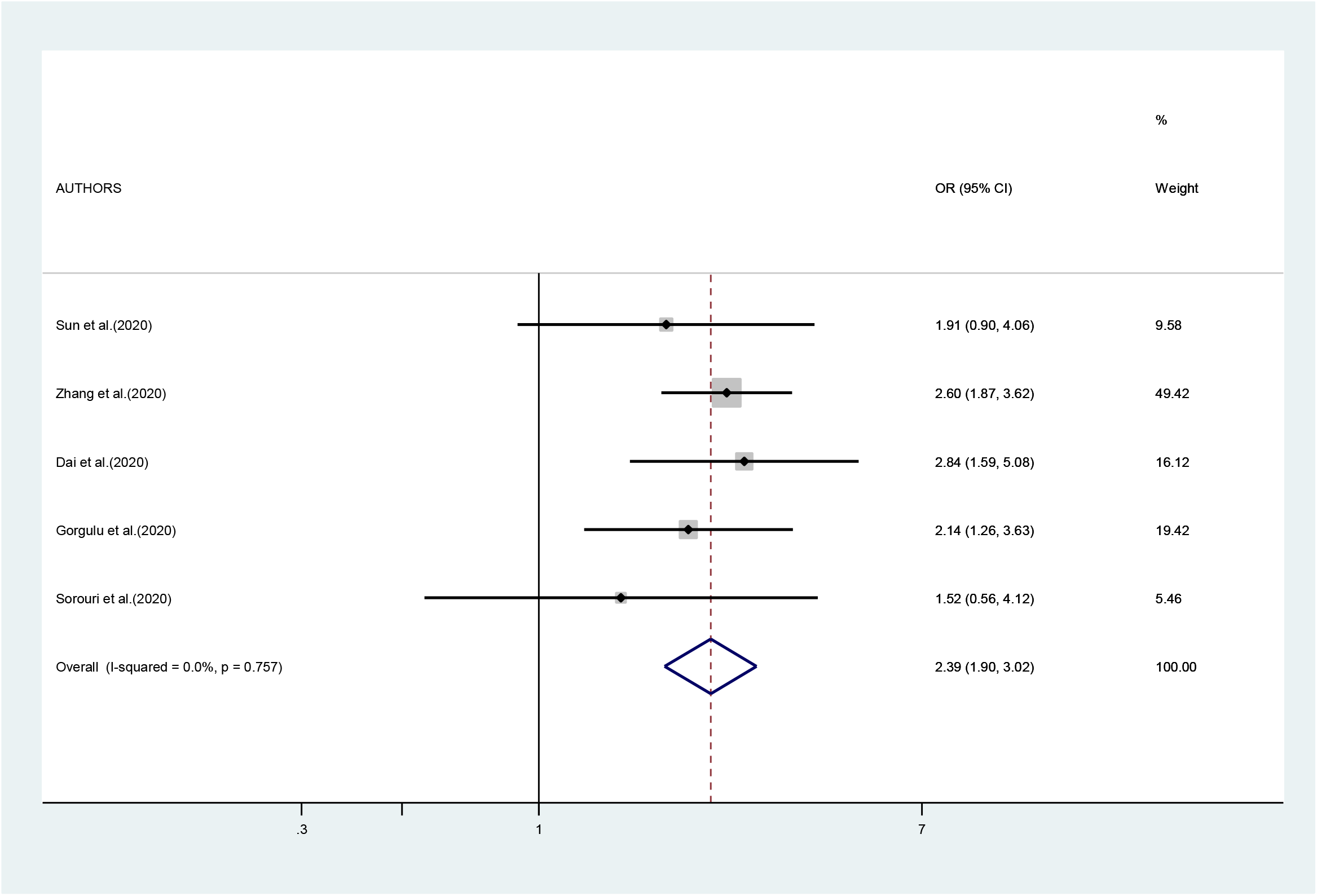
All type of cancer: Outcome 2 (access to ICU)

**Fig 4.**
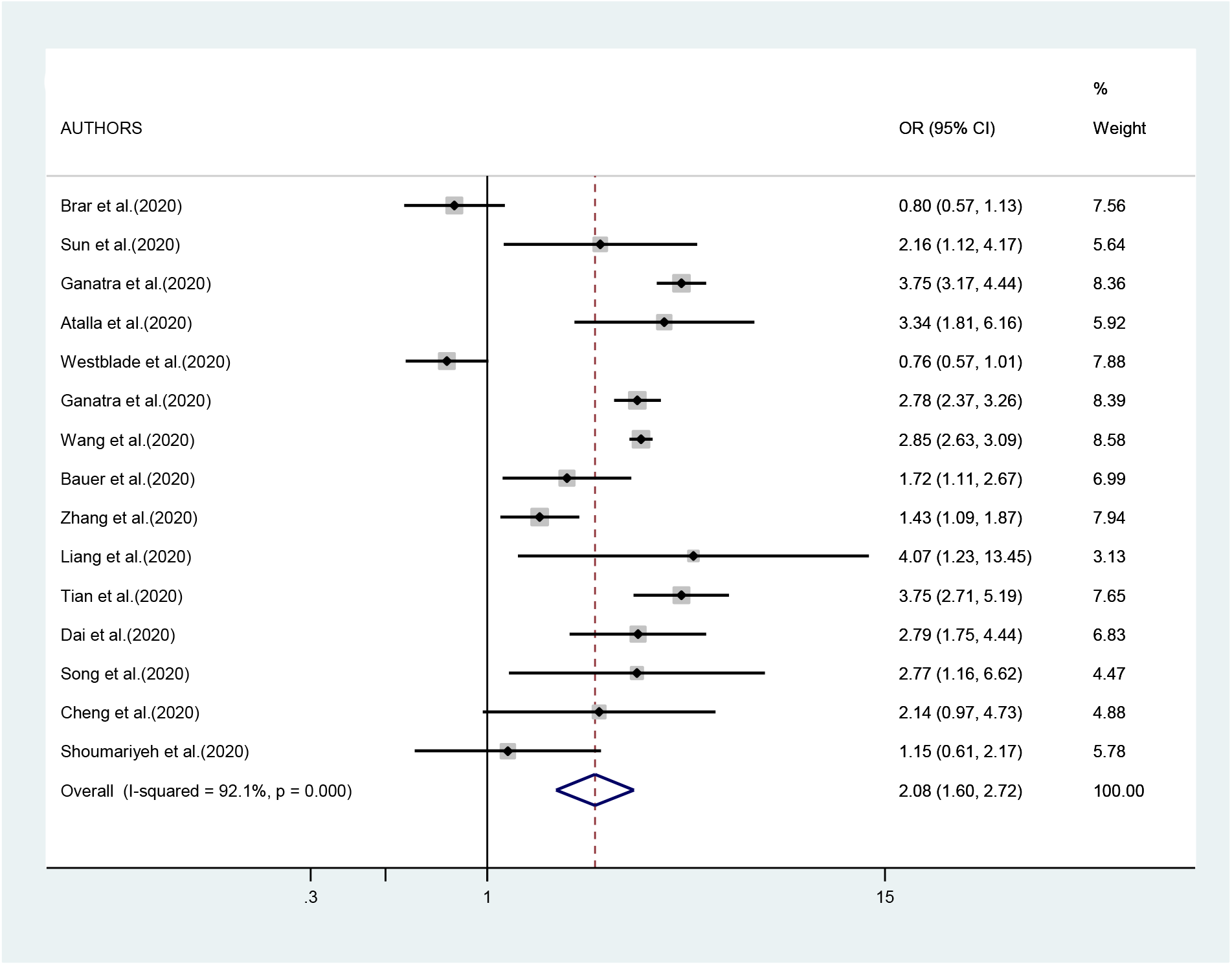
All type of cancer: Outcome 3 o 4 (Hospitalization or Severity of symptoms)

In the analysis by geographic region (Figure 2), the association between SARS-CoV-2 infection and mortality in cancer patients was stronger, and less heterogeneous, in studies from Asia (OR 2.92; 95% CI 2.13-4.01, I^2^ 37.8%) than in studies from either Europe (OR 2.37; 95% CI 1.65-3.40; I^2^ 67.9%) or North America (OR 1.97; 95% CI 1.31-2.97; I^2^ 95.9%). Too few studies were available on the other outcomes to justify a meta-analysis stratified by region of origin.

The cumulative meta-analysis, based on date of publication of subsequent studies of mortality (all type of cancer), showed a stronger association in the studies published before July 2020 than in studies published later.

As shown in Supplementary Figure 4, we found no evidence of publication bias in the meta-analysis concerning mortality (p-value of Egger’s test 0.67). The number of studies included in the other meta-analyses was too low to yield meaningful results on publication bias.

In the sensitivity analysis based on QA, the pooled OR of mortality results of studies with acceptable quality was not different from that of results of good-quality studies: OR 2.25 (95% CI 1.73-2.94) vs. OR 2.50 (95% CI 1.47-4.26). When we repeated the analysis after excluding one study at a time, we did not identify a major effect of any single study; in particular, the exclusion of the only study that suggested a negative association between SARS-CoV-2 infection and mortality [18] yielded a pooled OR of 2.41 (95% CI 1.95-2,99, I^2^ 85.5%). The association with mortality was less pronounced in studies whose results were reported by the authors (OR 2.11; 95% CI 1.55-2.87) compared to studies whose results were calculated by us (OR 2.66; 95% CI 1.97-3.60%), although the difference was not statistically significant. [Supplementary Figure 3]

Figure 5 presents the results of the meta-analysis of eight studies on mortality in patients with hematologic neoplasms. The pooled RR was 2.14 (95% CI 1.87-2.44, I^2^ 20.8%). Results for other outcomes (admission to ICU, hospitalization, severity of symptoms) were too sparse to conduct a meta-analysis.

**Fig 5.**
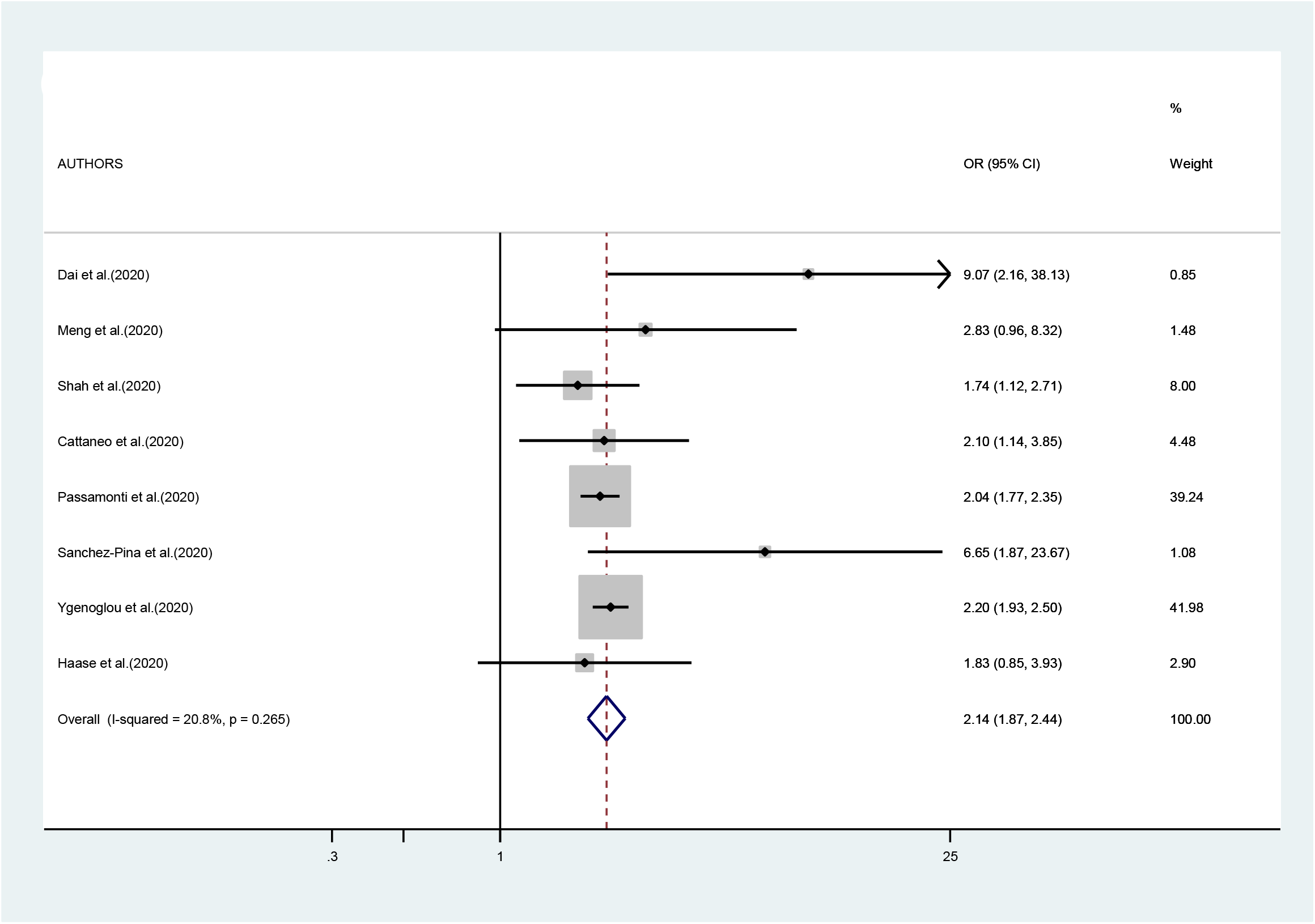
Hematological tumors: Outcome 1 (Mortality)

**Fig 6.**
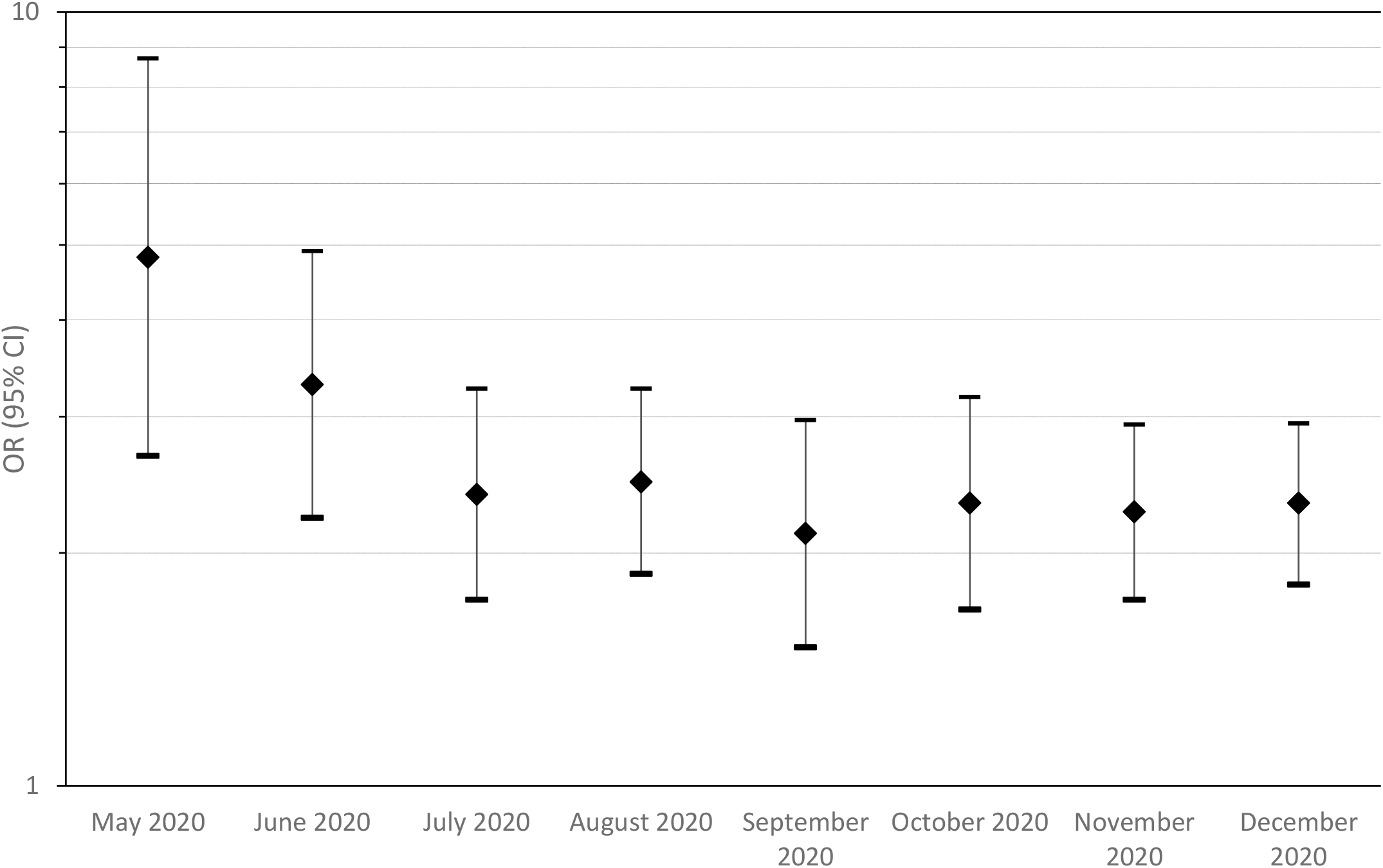
Meta-analysis of studies of mortality (all type of cancer) by month of publication

## Discussion

Since the beginning of the SARS-CoV-2 pandemic, cancer patients affected by COVID-19 have been identified to be at increased risk of poor prognosis, together with other vulnerable categories of patients as those affected by cardiovascular disease, diabetes, kidney injury, obesity, or stroke [46]. However, how much SARS-CoV-2 infection actually resulted in more severe outcomes in cancer patients compared to other patients and what caused their worse clinical course has not been fully clarified. In a previous editorial, some of us addressed the issue of the different interactions that COVID-19 and cancer may have [47]. On one hand, it is interesting to study how COVID-19 evolves in patients with cancer, by assessing whether the infection in these patients has a more severe course than in a control group affected by the infection but without cancer. On the other hand, it is important to identify the effects that the pandemic itself has determined in patients with cancer, including reduced access to treatment, delay in diagnosis for postponed screening, increased time between follow-up visits, and change in treatment organization. Acquiring more severe infection could be due to both components.

In this systematic review and meta-analysis we focused on the effect that SARS-CoV-2 infection had in patients with cancer compared with those without cancer in terms of mortality, ICU access, and severity of COVID-19 (hospitalization or severity of symptoms). We found that patients with cancer and SARS-CoV-2 infection have a two-fold higher risk of experiencing these adverse outcomes compared to non-cancer patients. Our results are in agreement with those of the meta-analysis by Venkatesulu et al. [48], who included a smaller number of studies, mainly from China, and reported an OR of 2.54 (95% CI 1.47-4.42) for mortality in cancer patients with concurrent COVID-19, compared to non-cancer patients. Similar to our results, these authors also reported a stronger association in studies from China than in those from other regions. Our results are also similar to those by Zhang et al. [49], who reported a meta-analysis of five studies from China, yielding a meta-OR of 2.63, with limited heterogeneity; compared to these early reports, we included more studies in our meta-analysis, which should lead to a more robust and precise risk estimate.

The higher risk of mortality in studies from China compared to those from other countries could be explained by the fact that some of the studies from China were conducted during the very early phase of the infection, when diagnosis and treatment for SARS-CoV-2 might have been delayed, resulting in higher death rate. This interpretation is reinforced by the results of the cumulative meta-analysis, that showed a stronger effect detected in the early studies compared to later studies.

Our summary results on risk of ICU admission and severity of COVID-19 indicated a somehow weaker association than that reported by other authors. An early meta-analysis reported a 3-fold increase for ICU admission, an almost 4-fold increase for a SARS-CoV-2 infection classified as severe, and a 5-fold increase in being intubated [51]. The fact that our values are lower might be explained by the inclusion of studies conducted when management of cancer patients with SARS-CoV-2 infection was more effective.

Immunosuppression and impaired T-cell response due to therapies may underlie the worse outcome in hematologic cancer diseases, even if some authors suggested that the attenuated inflammatory response in hematological patients can protect from severe COVID-19 morbidity [52]. The results of our meta-analysis confirm a higher mortality from COVID in patients with haematological neoplasms compared to non-neoplastic patients, with limited heterogeneity, with a pooled risk estimate similar to that for all cancers combined.

We were not able to derive pooled results for other specific cancers. Results for patients with hematologic and solid neoplasms were compared in some individual studies. In particular, Desai et al. [53] reported a higher mortality in the former group, but the comparison was not adjusted for age and type of therapy.

Although our study provides the most precise measure to date of the effect of COVID-19 in cancer patients, it suffers from some limitations. Many studies included in our analysis did not provide results adjusted for important determinants such as sex, age, comorbidities, and therapy. As mentioned above, we were not able to analyse specific cancers other than hematologic neoplasms, because results were too sparse.

In conclusion our meta-analysis confirms, by giving a more precise and accurate estimation, evidence to the hypothesis of an association between all type of cancer (and more specific hematologic neoplasm) and a worst outcome on Mortality, ICU admission and Severity of COVID-19.

Future studies will be able to better analyse this association for the different subtypes of cancer too. Furthermore, they will eventually be able to evaluate whether the difference among vaccinated population is reduced.

## Data Availability

All the primary data are available from the first Author

## Declarations

### Availability of data and material

all the primary data are available from the first Author.

### Code availability

all the software code is available from the first Author.

### Author’s contributions

- PB, GDF and GV designed the study
- GDF, GV, FT, MA identified studies and extracted the data
- GDF, GV and PB conducted statistical analysis and drafted the manuscript
- All Authors reviewed the manuscript

## Supplementary Figures

**Fig S1.**
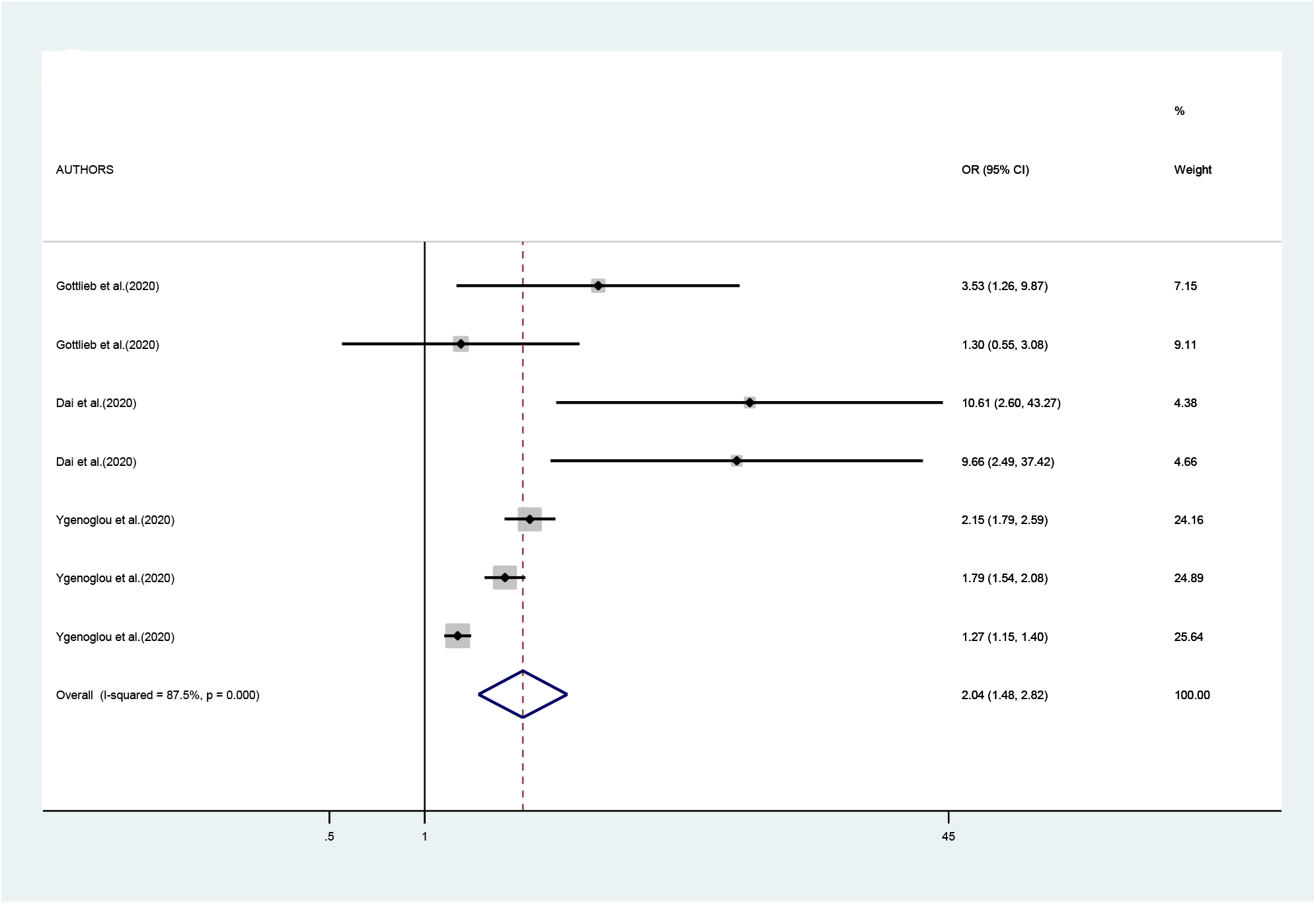
Hematological tumors: Outcome 2 (access to ICU), 3 (Hospitalization), 4 (Severity of Symptoms)

**FigS2.**
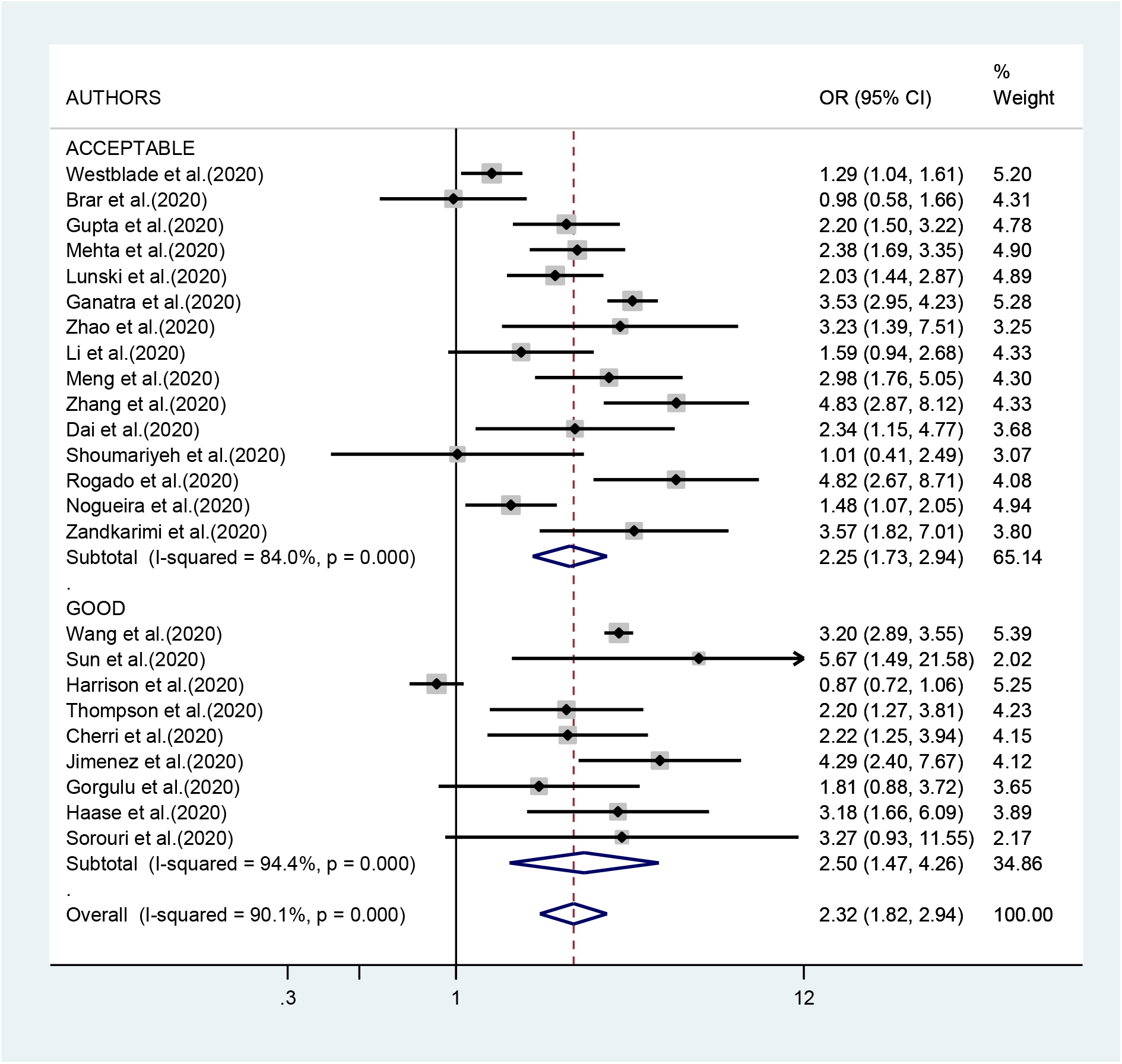
Quality Assestment by CASP_all type of cancer Outcome 1 (Mortality): Acceptable studies (CASP>6-9.5) vs Good studies (CASP>9.5-12)

**Fig S3.**
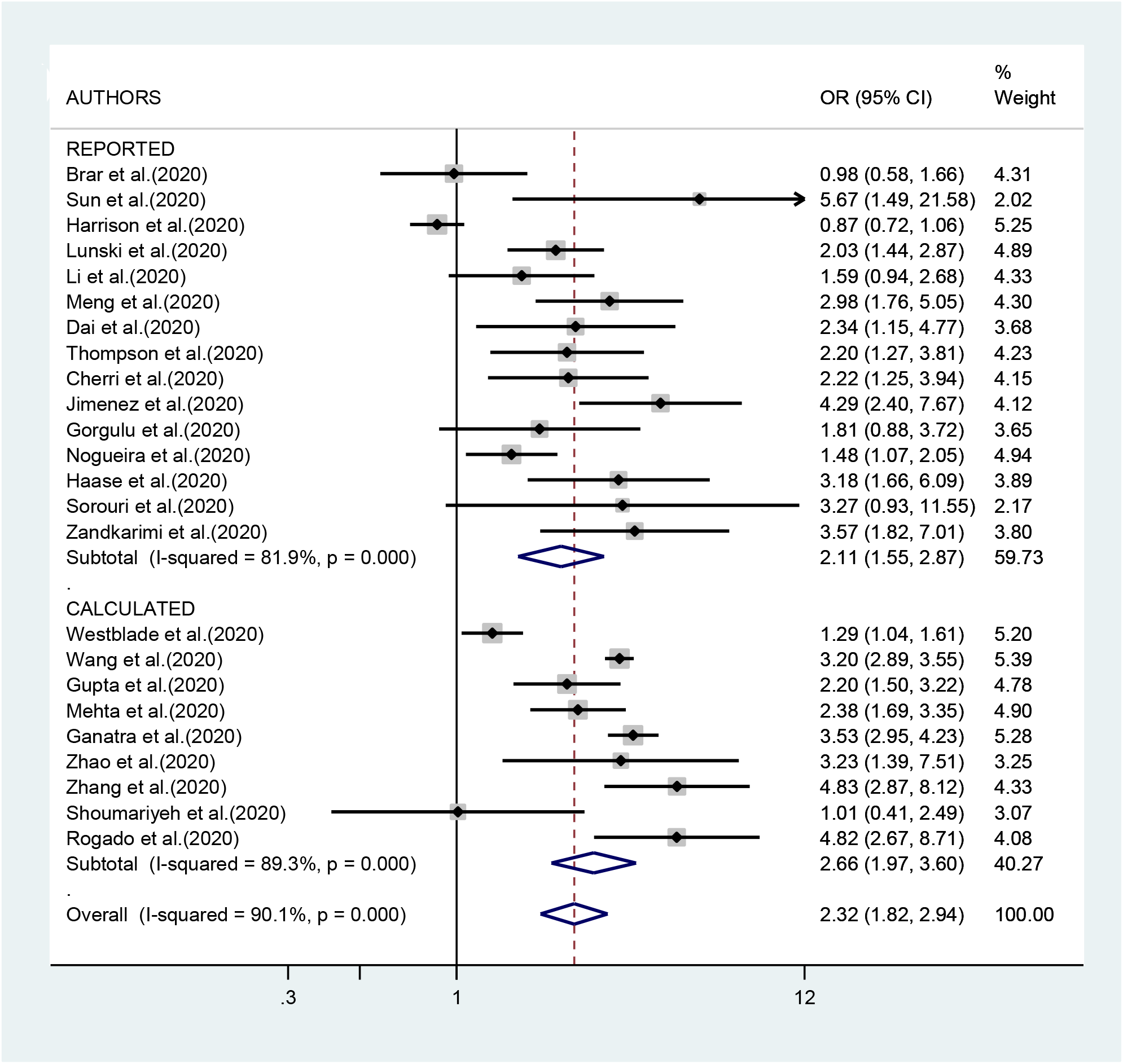
All type of cancer: Outcome 1 (Mortality) Reported vs Calculated OR

**Fig S4.**
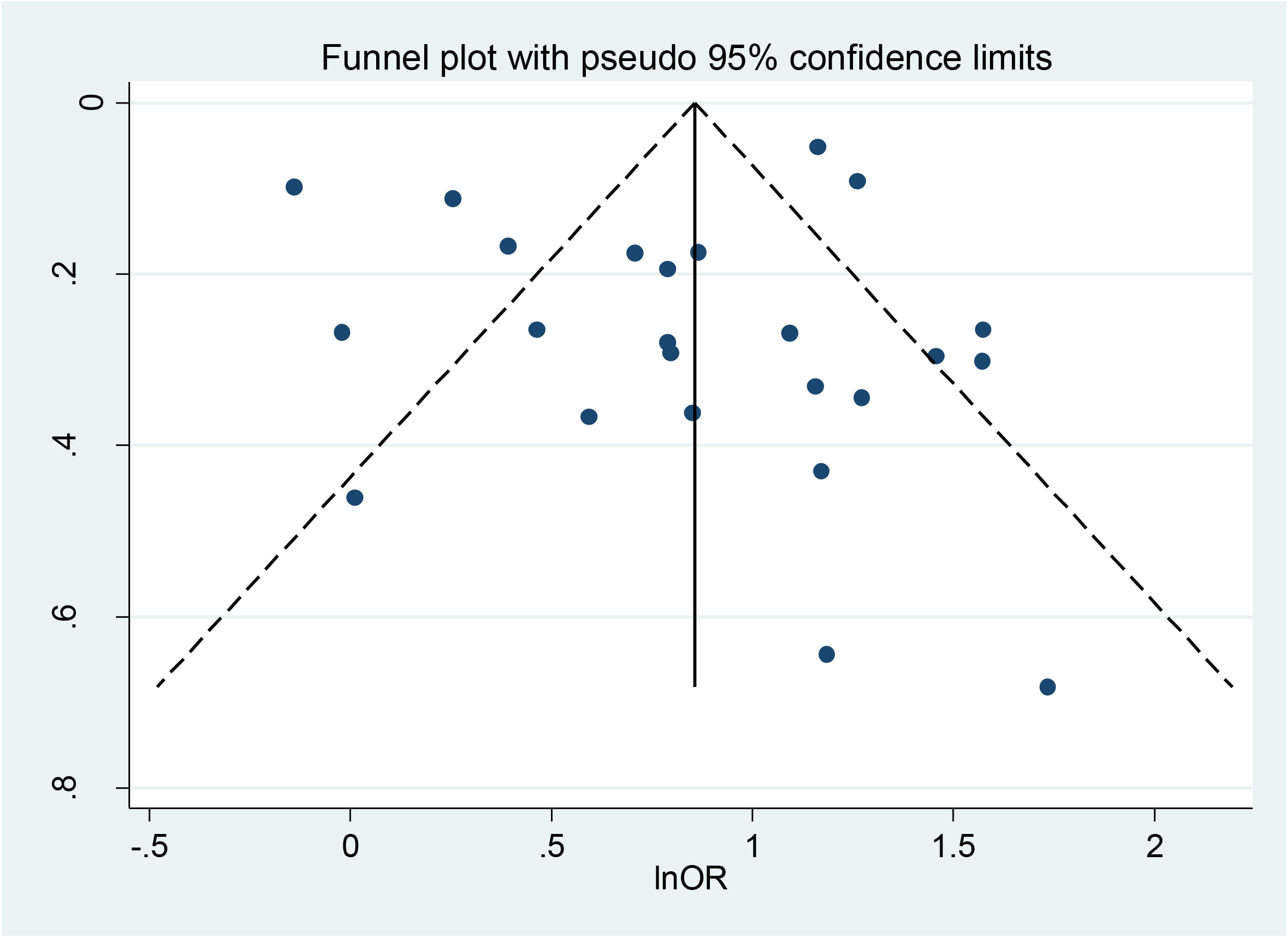
Funnel Plot: All type of cancer Outcome1 (Mortality) p-value of Egger’s test: P= 0.665 for Mortality studies

## References

1. X. Fang et al., «Epidemiological, comorbidity factors with severity and prognosis of COVID-19: a systematic review and meta-analysis», Aging, vol. 12, n. 13, pagg. 12493–12503, lug. 2020, doi: 10.18632/aging.103579

2. M. Kamboje K. A. Sepkowitz, «Nosocomial infections in patients with cancer», Lancet Oncol., vol. 10, n. 6, pagg. 589–597, giu. 2009, doi: 10.1016/S1470-2045(09)70069-5

3. M. M. Rüthrich et al., «COVID-19 in cancer patients: clinical characteristics and outcome-an analysis of the LEOSS registry», Ann. Hematol., vol. 100, n. 2, pagg. 383–393, feb. 2021, doi: 10.1007/s00277-020-04328-4

4. L. Y. W. Lee et al., «COVID-19 prevalence and mortality in patients with cancer and the effect of primary tumour subtype and patient demographics: a prospective cohort study», Lancet Oncol., vol. 21, n. 10, pagg. 1309–1316, ott. 2020, doi: 10.1016/S1470-2045(20)30442-3

5. Moher D on PROSPERO registry, Liberati A, Tetzlaff J, Altman DG; PRISMA Group. Preferred reporting items for systematic reviews and meta-analyses: the PRISMA statement. BMJ. 2009;339:b2535

6. Critical Appraisal Skills Programme (2018a). CASP Cohort Study Checklist. [online] Available at: https://casp-uk.net/casp-tools-checklists/. Accessed: 27 March 2019

7. DerSimonian R, Laird N. Meta-analysis in clinical trials. Control Clin Trials. 1986;7:177–88

8. Higgins JP, Thompson SG. Quantifying heterogeneity in a meta-analysis. Stat Med. 2002 Jun 15;21(11):1539–58. doi: 10.1002/sim.1186. PMID: 12111919.

9. Egger M, Davey Smith G, Schneider M, Minder C. Bias in meta-analysis detected by a simple, graphical test. BMJ. 1997;315:629–34

10. StataCorp. 2019. Stata Statistical Software: Release 16. College Station, TX: StataCorp LLC.

11. Dai M, Liu D, Liu M, Zhou F, Li G, Chen Z, Zhang Z, You H, Wu M, Zheng Q, Xiong Y, Xiong H, Wang C, Chen C, Xiong F, Zhang Y, Peng Y, Ge S, Zhen B, Yu T, Wang L, Wang H, Liu Y, Chen Y, Mei J, Gao X, Li Z, Gan L, He C, Li Z, Shi Y, Qi Y, Yang J, Tenen DG, Chai L, Mucci LA, Santillana M, Cai H. Patients with Cancer Appear More Vulnerable to SARS-CoV-2: A Multicenter Study during the COVID-19 Outbreak. Cancer Discov. 2020 Jun;10(6):783–791. doi: 10.1158/2159-8290.CD-20-0422. Epub 2020 Apr 28. PMID: 32345594; PMCID: PMC7309152.

12. Haase N, Plovsing R, Christensen S, Poulsen LM, Brøchner AC, Rasmussen BS, Helleberg M, Jensen JUS, Andersen LPK, Siegel H, Ibsen M, Jørgensen V, Winding R, Iversen S, Pedersen HP, Madsen J, Sølling C, Garcia RS, Michelsen J, Mohr T, Mannering A, Espelund US, Bundgaard H, Kirkegaard L, Smitt M, Buck DL, Ribergaard NE, Pedersen HS, Christensen BV, Perner A. Characteristics, interventions, and longer term outcomes of COVID-19 ICU patients in Denmark-A nationwide, observational study. Acta Anaesthesiol Scand. 2021 Jan;65(1):68–75. doi: 10.1111/aas.13701. Epub 2020 Oct 3. PMID: 32929715.

13. Meng Y, Lu W, Guo E, Liu J, Yang B, Wu P, Lin S, Peng T, Fu Y, Li F, Wang Z, Li Y, Xiao R, Liu C, Huang Y, Lu F, Wu X, You L, Ma D, Sun C, Wu P, Chen G. Cancer history is an independent risk factor for mortality in hospitalized COVID-19 patients: a propensity score-matched analysis. J Hematol Oncol. 2020 Jun 10;13(1):75. doi: 10.1186/s13045-020-00907-0. PMID: 32522278; PMCID: PMC7286218.

14. Sun L, Surya S, Le AN, Desai H, Doucette A, Gabriel P, Ritchie M, Rader D, Maillard I, Bange E, Huang A, Vonderheide RH, DeMichele A, Verma A, Mamtani R, Maxwell KN. Rates of COVID-19-related Outcomes in Cancer compared to non-Cancer Patients. medRxiv [Preprint]. 2020 Aug 15:2020.08.14.20174961. doi: 10.1101/2020.08.14.20174961. Update in: JNCI Cancer Spectr. 2021 Jan 21;5(1):pkaa120. PMID: 32817956; PMCID: PMC7430598.

15. Nogueira PJ, de Araújo Nobre M, Costa A, Ribeiro RM, Furtado C, Bacelar Nicolau L, Camarinha C, Luís M, Abrantes R, Vaz Carneiro A. The Role of Health Preconditions on COVID-19 Deaths in Portugal: Evidence from Surveillance Data of the First 20293 Infection Cases. J Clin Med. 2020 Jul 24;9(8):2368. doi: 10.3390/jcm9082368. PMID: 32722159; PMCID: PMC7464004.

16. Zandkarimi E, Moradi G, Mohsenpour B. The Prognostic Factors Affecting the Survival of Kurdistan Province COVID-19 Patients: A Cross-sectional Study From February to May 2020. Int J Health Policy Manag. 2020 Aug 22. doi: 10.34172/ijhpm.2020.155. Epub ahead of print. PMID: 32861230.

17. Zhao Y, Nie HX, Hu K, Wu XJ, Zhang YT, Wang MM, Wang T, Zheng ZS, Li XC, Zeng SL. Abnormal immunity of non-survivors with COVID-19: predictors for mortality. Infect Dis Poverty. 2020 Aug 3;9(1):108. doi: 10.1186/s40249-020-00723-1. PMID: 32746940; PMCID: PMC7396941.

18. Harrison SL, Fazio-Eynullayeva E, Lane DA, Underhill P, Lip GYH. Comorbidities associated with mortality in 31,461 adults with COVID-19 in the United States: A federated electronic medical record analysis. PLoS Med. 2020 Sep 10;17(9):e1003321. doi: 10.1371/journal.pmed.1003321. PMID: 32911500; PMCID: PMC7482833.

19. Gupta S, Hayek SS, Wang W, Chan L, Mathews KS, Melamed ML, Brenner SK, Leonberg-Yoo A, Schenck EJ, Radbel J, Reiser J, Bansal A, Srivastava A, Zhou Y, Sutherland A, Green A, Shehata AM, Goyal N, Vijayan A, Velez JCQ, Shaefi S, Parikh CR, Arunthamakun J, Athavale AM, Friedman AN, Short SAP, Kibbelaar ZA, Abu Omar S, Admon AJ, Donnelly JP, Gershengorn HB, Hernán MA, Semler MW, Leaf DE; STOP-COVID Investigators. Factors Associated With Death in Critically Ill Patients With Coronavirus Disease 2019 in the US. JAMA Intern Med. 2020 Nov 1;180(11):1436–1447. doi: 10.1001/jamainternmed.2020.3596. Erratum in: JAMA Intern Med. 2020 Nov 1;180(11):1555. Erratum in: JAMA Intern Med. 2021 Aug 1;181(8):1144. PMID: 32667668; PMCID: PMC7364338.

20. Ganatra S, Dani SS, Redd R, Rieger-Christ K, Patel R, Parikh R, Asnani A, Bang V, Shreyder K, Brar SS, Singh A, Kazi DS, Guha A, Hayek SS, Barac A, Gunturu KS, Zarwan C, Mosenthal AC, Yunus SA, Kumar A, Patel JM, Patten RD, Venesy DM, Shah SP, Resnic FS, Nohria A, Baron SJ. Outcomes of COVID-19 in Patients With a History of Cancer and Comorbid Cardiovascular Disease. J Natl Compr Canc Netw. 2020 Nov 3:1–10. doi: 10.6004/jnccn.2020.7658. Epub ahead of print. PMID: 33142266.

21. Westblade LF, Brar G, Pinheiro LC, Paidoussis D, Rajan M, Martin P, Goyal P, Sepulveda JL, Zhang L, George G, Liu D, Whittier S, Plate M, Small CB, Rand JH, Cushing MM, Walsh TJ, Cooke J, Safford MM, Loda M, Satlin MJ. SARS-CoV-2 Viral Load Predicts Mortality in Patients with and without Cancer Who Are Hospitalized with COVID-19. Cancer Cell. 2020 Nov 9;38(5):661–671.e2. doi: 10.1016/j.ccell.2020.09.007. Epub 2020 Sep 15. PMID: 32997958; PMCID: PMC7492074.

22. Wang B, Van Oekelen O, Mouhieddine TH, Del Valle DM, Richter J, Cho HJ, Richard S, Chari A, Gnjatic S, Merad M, Jagannath S, Parekh S, Madduri D. A tertiary center experience of multiple myeloma patients with COVID-19: lessons learned and the path forward. J Hematol Oncol. 2020 Jul 14;13(1):94. doi: 10.1186/s13045-020-00934-x. PMID: 32664919; PMCID: PMC7359431.

23. Cherri S, Lemmers DHL, Noventa S, Abu Hilal M, Zaniboni A. Outcome of oncological patients admitted with COVID-19: experience of a hospital center in northern Italy. Ther Adv Med Oncol. 2020 Sep 30;12:1758835920962370. doi: 10.1177/1758835920962370. PMID: 33062065; PMCID: PMC7533930.

24. Görgülü Ö, Duyan M. Effects of Comorbid Factors on Prognosis of Three Different Geriatric Groups with COVID-19 Diagnosis. SN Compr Clin Med. 2020 Nov 18:1–12. doi: 10.1007/s42399-020-00645-x. Epub ahead of print. PMID: 33225222; PMCID: PMC7671936.

25. Jiménez E, Fontán-Vela M, Valencia J, Fernandez-Jimenez I, Álvaro-Alonso EA, Izquierdo-García E, Lazaro Cebas A, Gallego Ruiz-Elvira E, Troya J, Tebar-Martinez AJ, Garcia-Marina B, Peña-Lillo G, Abad-Motos A, Macaya L, Ryan P, Pérez-Butragueño M; COVID@HUIL Working Group; COVID@HUIL Working Group. Characteristics, complications and outcomes among 1549 patients hospitalised with COVID-19 in a secondary hospital in Madrid, Spain: a retrospective case series study. BMJ Open. 2020 Nov 10;10(11):e042398. doi: 10.1136/bmjopen-2020-042398. PMID: 33172949; PMCID: PMC7656887.

26. Thompson JV, Meghani NJ, Powell BM, Newell I, Craven R, Skilton G, Bagg LJ, Yaqoob I, Dixon MJ, Evans EJ, Kambele B, Rehman A, Ng Man Kwong G. Patient characteristics and predictors of mortality in 470 adults admitted to a district general hospital in England with Covid-19. Epidemiol Infect. 2020 Nov 24;148:e285. doi: 10.1017/S0950268820002873. PMID: 33228824; PMCID: PMC7729176.

27. Li Q, Chen L, Li Q, He W, Yu J, Chen L, Cao Y, Chen W, Di Wu, Dong F, Cai L, Ran Q, Li L, Liu Q, Ren W, Gao F, Wang H, Chen Z, Gale RP, Hu Y. Cancer increases risk of in-hospital death from COVID-19 in persons <65 years and those not in complete remission. Leukemia. 2020 Sep;34(9):2384–2391. doi: 10.1038/s41375-020-0986-7. Epub 2020 Jul 20. PMID: 32690880; PMCID: PMC7371786.

28. Shoumariyeh K, Biavasco F, Ihorst G, Rieg S, Nieters A, Kern WV, Miething C, Duyster J, Engelhardt M, Bertz H. Covid-19 in patients with hematological and solid cancers at a Comprehensive Cancer Center in Germany. Cancer Med. 2020 Nov;9(22):8412–8422. doi: 10.1002/cam4.3460. Epub 2020 Sep 15. PMID: 32931637; PMCID: PMC7666742.

29. Mehta V, Goel S, Kabarriti R, Cole D, Goldfinger M, Acuna-Villaorduna A, Pradhan K, Thota R, Reissman S, Sparano JA, Gartrell BA, Smith RV, Ohri N, Garg M, Racine AD, Kalnicki S, Perez-Soler R, Halmos B, Verma A. Case Fatality Rate of Cancer Patients with COVID-19 in a New York Hospital System. Cancer Discov. 2020 Jul;10(7):935–941. doi: 10.1158/2159-8290.CD-20-0516. Epub 2020 May 1. PMID: 32357994; PMCID: PMC7334098.

30. Rogado J, Obispo B, Pangua C, Serrano-Montero G, Martín Marino A, Pérez-Pérez M, López-Alfonso A, Gullón P, Lara MÁ. Covid-19 transmission, outcome and associated risk factors in cancer patients at the first month of the pandemic in a Spanish hospital in Madrid. Clin Transl Oncol. 2020 Dec;22(12):2364–2368. doi: 10.1007/s12094-020-02381-z. Epub 2020 May 25. PMID: 32449128; PMCID: PMC7246222.

31. Brar G, Pinheiro LC, Shusterman M, Swed B, Reshetnyak E, Soroka O, Chen F, Yamshon S, Vaughn J, Martin P, Paul D, Hidalgo M, Shah MA. COVID-19 Severity and Outcomes in Patients With Cancer: A Matched Cohort Study. J Clin Oncol. 2020 Nov 20;38(33):3914–3924. doi: 10.1200/JCO.20.01580. Epub 2020 Sep 28. PMID: 32986528; PMCID: PMC7676890.

32. Zhang B, Yu Y, Hubert SM, Zhang Y, Lu J, Liu S, Xie F, Zhao L, Lei X, Deng W, Chen J, Li Y. Prognostic Value of Pro-Inflammatory Neutrophils and C-Reactive Protein in Cancer Patient With Coronavirus Disease 2019: A Multi-Center, Retrospective Study. Front Pharmacol. 2020 Oct 22;11:576994. doi: 10.3389/fphar.2020.576994. PMID: 33192519; PMCID: PMC7656194.

33. Sorouri M, Kasaeian A, Mojtabavi H, Radmard AR, Kolahdoozan S, Anushiravani A, Khosravi B, Pourabbas SM, Eslahi M, Sirusbakht A, Khodabakhshi M, Motamedi F, Azizi F, Ghanbari R, Rajabi Z, Sima AR, Rad S, Abdollahi M. Clinical characteristics, outcomes, and risk factors for mortality in hospitalized patients with COVID-19 and cancer history: a propensity score-matched study. Infect Agent Cancer. 2020 Dec 17;15(1):74. doi: 10.1186/s13027-020-00339-y. PMID: 33334375; PMCID: PMC7745169.

34. Lunski MJ, Burton J, Tawagi K, Maslov D, Simenson V, Barr D, Yuan H, Johnson D, Matrana M, Cole J, Larned Z, Moore B. Multivariate mortality analyses in COVID-19: Comparing patients with cancer and patients without cancer in Louisiana. Cancer. 2021 Jan 15;127(2):266–274. doi: 10.1002/cncr.33243. Epub 2020 Oct 28. PMID: 33112411.

35. Atalla E, Kalligeros M, Giampaolo G, Mylona EK, Shehadeh F, Mylonakis E. Readmissions among patients with COVID-19. Int J Clin Pract. 2021 Mar;75(3):e13700. doi: 10.1111/ijcp.13700. Epub 2020 Oct 12. PMID: 32894801.

36. Cheng WT, Ke YH, Yang GY, Sun H, Chen Y, Ying RY, Zeng XH, Shen D, Tang KJ, Xu K, Yu F. Analysis of clinical features of COVID-19 in cancer patients. Acta Oncol. 2020 Nov;59(11):1393–1396. doi: 10.1080/0284186X.2020.1810313. Epub 2020 Aug 28. PMID: 32857662.

37. Song J, Zeng M, Wang H, Qin C, Hou HY, Sun ZY, Xu SP, Wang GP, Guo CL, Deng YK, Wang ZC, Ma J, Pan L, Liao B, Du ZH, Feng QM, Liu Y, Xie JG, Liu Z. Distinct effects of asthma and COPD comorbidity on disease expression and outcome in patients with COVID-19. Allergy. 2021 Feb;76(2):483–496. doi: 10.1111/all.14517. Epub 2020 Aug 11. PMID: 32716553.

38. Liang W, Liang H, Ou L, Chen B, Chen A, Li C, Li Y, Guan W, Sang L, Lu J, Xu Y, Chen G, Guo H, Guo J, Chen Z, Zhao Y, Li S, Zhang N, Zhong N, He J; China Medical Treatment Expert Group for COVID-19. Development and Validation of a Clinical Risk Score to Predict the Occurrence of Critical Illness in Hospitalized Patients With COVID-19. JAMA Intern Med. 2020 Aug 1;180(8):1081–1089. doi: 10.1001/jamainternmed.2020.2033. PMID: 32396163; PMCID: PMC7218676.

39. Bauer AZ, Gore R, Sama SR, Rosiello R, Garber L, Sundaresan D, McDonald A, Arruda P, Kriebel D. Hypertension, medications, and risk of severe COVID-19: A Massachusetts community-based observational study. J Clin Hypertens (Greenwich). 2021 Jan;23(1):21–27. doi: 10.1111/jch.14101. Epub 2020 Nov 21. PMID: 33220171; PMCID: PMC7753489.

40. Tian J, Yuan X, Xiao J, Zhong Q, Yang C, Liu B, Cai Y, Lu Z, Wang J, Wang Y, Liu S, Cheng B, Wang J, Zhang M, Wang L, Niu S, Yao Z, Deng X, Zhou F, Wei W, Li Q, Chen X, Chen W, Yang Q, Wu S, Fan J, Shu B, Hu Z, Wang S, Yang XP, Liu W, Miao X, Wang Z. Clinical characteristics and risk factors associated with COVID-19 disease severity in patients with cancer in Wuhan, China: a multicentre, retrospective, cohort study. Lancet Oncol. 2020 Jul;21(7):893–903. doi: 10.1016/S1470-2045(20)30309-0. Epub 2020 May 29. PMID: 32479790; PMCID: PMC7259911.

41. Yigenoglu TN, Ata N, Altuntas F, Bascı S, Dal MS, Korkmaz S, Namdaroglu S, Basturk A, Hacıbekiroglu T, Dogu MH, Berber İ, Dal K, Erkurt MA, Turgut B, Ulgu MM, Celik O, Imrat E, Birinci S. The outcome of COVID-19 in patients with hematological malignancy. J Med Virol. 2021 Feb;93(2):1099–1104. doi: 10.1002/jmv.26404. Epub 2020 Aug 26. PMID: 32776581; PMCID: PMC7436524.

42. Shah V, Ko Ko T, Zuckerman M, Vidler J, Sharif S, Mehra V, Gandhi S, Kuhnl A, Yallop D, Avenoso D, Rice C, Sanderson R, Sarma A, Marsh J, de Lavallade H, Krishnamurthy P, Patten P, Benjamin R, Potter V, Ceesay MM, Mufti GJ, Norton S, Pagliuca A, Galloway J, Kulasekararaj AG. Poor outcome and prolonged persistence of SARS-CoV-2 RNA in COVID-19 patients with haematological malignancies; King’s College Hospital experience. Br J Haematol. 2020 Sep;190(5):e279–e282. doi: 10.1111/bjh.16935. Epub 2020 Aug 10. PMID: 32526039; PMCID: PMC7307054.

43. Sanchez-Pina JM, Rodríguez Rodriguez M, Castro Quismondo N, Gil Manso R, Colmenares R, Gil Alos D, Paciello ML, Zafra D, Garcia-Sanchez C, Villegas C, Cuellar C, Carreño-Tarragona G, Zamanillo I, Poza M, Iñiguez R, Gutierrez X, Alonso R, Rodríguez A, Folgueira MD, Delgado R, Ferrari JM, Lizasoain M, Aguado JM, Ayala R, Martinez-Lopez J, Calbacho M. Clinical course and risk factors for mortality from COVID-19 in patients with haematological malignancies. Eur J Haematol. 2020 Nov;105(5):597–607. doi: 10.1111/ejh.13493. Epub 2020 Aug 11. PMID: 32710500.

44. Passamonti F, Cattaneo C, Arcaini L, Bruna R, Cavo M, Merli F, Angelucci E, Krampera M, Cairoli R, Della Porta MG, Fracchiolla N, Ladetto M, Gambacorti Passerini C, Salvini M, Marchetti M, Lemoli R, Molteni A, Busca A, Cuneo A, Romano A, Giuliani N, Galimberti S, Corso A, Morotti A, Falini B, Billio A, Gherlinzoni F, Visani G, Tisi MC, Tafuri A, Tosi P, Lanza F, Massaia M, Turrini M, Ferrara F, Gurrieri C, Vallisa D, Martelli M, Derenzini E, Guarini A, Conconi A, Cuccaro A, Cudillo L, Russo D, Ciambelli F, Scattolin AM, Luppi M, Selleri C, Ortu La Barbera E, Ferrandina C, Di Renzo N, Olivieri A, Bocchia M, Gentile M, Marchesi F, Musto P, Federici AB, Candoni A, Venditti A, Fava C, Pinto A, Galieni P, Rigacci L, Armiento D, Pane F, Oberti M, Zappasodi P, Visco C, Franchi M, Grossi PA, Bertù L, Corrao G, Pagano L, Corradini P; ITA-HEMA-COV Investigators. Clinical characteristics and risk factors associated with COVID-19 severity in patients with haematological malignancies in Italy: a retrospective, multicentre, cohort study. Lancet Haematol. 2020 Oct;7(10):e737–e745. doi: 10.1016/S2352-3026(20)30251-9. Epub 2020 Aug 13. PMID: 32798473; PMCID: PMC7426107.

45. Cattaneo C, Daffini R, Pagani C, Salvetti M, Mancini V, Borlenghi E, D’Adda M, Oberti M, Paini A, De Ciuceis C, Barbullushi K, Cancelli V, Belotti A, Re A, Motta M, Peli A, Bianchetti N, Anastasia A, Dalceggio D, Roccaro AM, Tucci A, Cairoli R, Muiesan ML, Rossi G. Clinical characteristics and risk factors for mortality in hematologic patients affected by COVID-19. Cancer. 2020 Dec 1;126(23):5069–5076. doi: 10.1002/cncr.33160. Epub 2020 Sep 10. PMID: 32910456.

46. Hu Y, Sun J, Dai Z, Deng H, Li X, Huang Q, Wu Y, Sun L, Xu Y. Prevalence and severity of corona virus disease 2019 (COVID-19): A systematic review and meta-analysis. J Clin Virol. 2020 Jun;127:104371. doi: 10.1016/j.jcv.2020.104371. Epub 2020 Apr 14

47. Hainault P and Boffetta P-SARS-Cov2, COVID19 and Cancer: three research questions casting a long shadow-Editorial for Current Opinion in Oncology – Cancer Biology section

48. Venkatesulu BP, Chandrasekar VT, Girdhar P, Advani P, Sharma A, Elumalai T, Hsieh C, Elghazawy HI, Verma V, Krishnan S. A systematic review and meta-analysis of cancer patients affected by a novel coronavirus. medRxiv [Preprint]. 2020 May 29:2020.05.27.20115303. doi: 10.1101/2020.05.27.20115303. PMID: 32511470; PMCID: PMC7265691

49. Zhang Y, Han H, Tian Y, Dong J, Yu Y, Kang Y, Xing L, Lian R, Zhang R, Xie D. Impact of cancer on mortality and severity of corona virus disease 2019: A protocol for systematic review and meta-analysis. Medicine (Baltimore). 2020 Oct 30;99(44):e23005

50. Patel R, Park J, Shah A, Saif MW. COVID-19 and Cancer Patients. Cancer Med J. 2020;3(1):40–48. Epub 2020 Apr

51. ElGohary GM, Hashmi S, Styczynski J, et al. The risk and prognosis of COVID-19 infection in cancer patients: A systematic review and meta-analysis [published online ahead of print, 2020 Jul 30]. Hematol Oncol Stem Cell Ther. 2020;S1658-3876(20)30122-9. doi:10.1016/j.hemonc.2020.07.005

52. Vijenthira A, Gong IY, Fox TA, Booth S, Cook G, Fattizzo B, Martín-Moro F, Razanamahery J, Riches JC, Zwicker J, Patell R, Vekemans MC, Scarfò L, Chatzikonstantinou T, Yildiz H, Lattenist R, Mantzaris I, Wood WA, Hicks LK. Outcomes of patients with hematologic malignancies and COVID-19: a systematic review and meta-analysis of 3377 patients. Blood. 2020 Dec 17;136(25):2881–2892

53. Desai, A, Gupta, R, Advani, S, Ouellette, L, Kuderer, NM, Lyman, GH, Li, A. Mortality in hospitalized patients with cancer and coronavirus disease 2019: A systematic review and meta-analysis of cohort studies. Cancer. 2021. https://doi.org/10.1002/cncr.33386

54. Hacisuleyman E, Hale C, Saito Y, Blachere NE, Bergh M, Conlon EG, Schaefer-Babajew DJ, DaSilva J, Muecksch F, Gaebler C, Lifton R, Nussenzweig MC, Hatziioannou T, Bieniasz PD, Darnell RB. Vaccine Breakthrough Infections with SARS-CoV-2 Variants. N Engl J Med. 2021 Jun 10;384(23):2212–2218. doi: 10.1056/NEJMoa2105000. Epub 2021 Apr 21. PMID: 33882219; PMCID: PMC8117968.

